# Performance of an automated anti-SARS-CoV-2 immunoassay in prepandemic cohorts

**DOI:** 10.1101/2020.08.07.20169987

**Authors:** Elena Riester, Beda Krieter, Peter Findeisen, Michael Laimighofer, Kathrin Schoenfeld, Tina Laengin, Christoph Niederhauser

## Abstract

**Background:** The Elecsys® Anti-SARS-CoV-2 immunoassay (Roche Diagnostics) was developed to provide an accurate and reliable method for the detection of antibodies to severe acute respiratory syndrome coronavirus 2 (SARS-CoV-2). We evaluated the specificity of the Elecsys Anti-SARS-CoV-2 immunoassay in prepandemic sample cohorts across five sites in Germany, Austria and Switzerland.

**Methods:** Specificity of the immunoassay was evaluated using anonymised, frozen, residual serum and/or plasma samples from blood donors or routine diagnostic testing. All samples were collected before September 2019 and therefore presumed negative for SARS-CoV-2-specific antibodies. Cohorts included samples from blood donors, pregnant women and paediatric patients. Point estimates and 95% confidence intervals (CIs) were calculated.

**Results:** Overall specificities for the Elecsys Anti-SARS-CoV-2 immunoassay in 9575 samples from blood donors (n = 6714) and diagnostic specimens (n = 2861) were 99.82% (95% CI 99.69-99.91) and 99.93% (95% CI 99.75-99.99), respectively. Among 2256 samples from pregnant women, specificity was 99.91% (95% CI 99.68-99.99). Among 205 paediatric samples, specificity was 100% (95% CI 98.22-100).

**Conclusion:** The Elecsys Anti-SARS-CoV-2 immunoassay demonstrated a very high specificity across blood donor samples and diagnostic specimens from Germany, Austria and Switzerland. Our findings support the use of the Elecsys Anti-SARS-CoV-2 immunoassay as a potential tool for determination of an immune response following previous exposure to SARS-CoV-2 in the general population, including in blood donors, pregnant women and paediatric populations.

## Introduction

The novel coronavirus, severe acute respiratory syndrome coronavirus 2 (SARS-CoV-2), is the causative agent in the Coronavirus Disease 2019 (COVID-19) outbreak, now deemed a pandemic, that first came to the world’s attention in December 2019.^1,2^ SARS-CoV-2 is an enveloped, singlestranded RNA virus that, like other coronaviruses, has a genome encoding 16 non-structural proteins and four structural proteins: spike (S), envelope (E), membrane (M) and nucleocapsid (N).^3,4^ It elicits an array of symptoms including fever, cough, breathing difficulties and fatigue. While most cases are non-severe, patients with severe disease may experience acute respiratory distress syndrome, and critically ill patients develop respiratory failure, septic shock and/or multiple organ dysfunction.^5^

Diagnosis of COVID-19 is usually based on direct detection of SARS-CoV-2 by nucleic acid amplification tests (NAATs) which indicate the presence of an acute infection.^6,7^ Following cessation of viral shedding, individuals who initially test negative for SARS-CoV-2 using NAATs may still test positive for the virus using antibody tests. Antibody tests identify individuals who have been exposed to the virus and developed an immune response, an approach that also allows assessment of the extent of exposure of a population.^8^ This information is of great interest for public health and is needed for decision-making regarding containment measures in given populations, as well as to support diagnosis, contact tracing and epidemiological studies.

Research into antibody responses against SARS-CoV-2 is rapidly developing and has begun to shed light on the timing of seroconversion, a critical consideration in serological testing. Evidence to date suggests that immunoglobulin M (IgM) antibodies are detectable within 5 days of symptom onset, immunoglobulin G (IgG) antibodies within 5-14 days,^9-11^ and immunoglobulin A (IgA) antibodies after approximately 3-6 days.^10,12^ The chronological order in which IgM and IgG antibodies develop appears to be highly variable, as are antibody levels.^11,13-15^ Together this supports the need for accurate serological tests that can detect both antibodies simultaneously.

The COVID-19 pandemic has prompted a rapid response including the launch of several different serological assays. Reliable information regarding the relative performance of these assays in a wide range of settings is urgently needed.

The Elecsys® Anti-SARS-CoV-2 serological assay (Roche Diagnostics International Ltd, Rotkreuz, Switzerland) is designed for in vitro qualitative detection of antibodies to SARS-CoV-2, including IgG, in human serum and plasma. Generally, the most abundant antibodies produced by the body are directed against the N protein.^16-19^ As such, this immunoassay is based on a modified recombinant protein representing the N antigen of SARS-CoV-2.

The clinical performance of the newly launched Elecsys Anti-SARS-CoV-2 serological assay is undergoing evaluation; however, this assay has demonstrated high sensitivity and specificity and acceptable agreement with EDI™ SARS-CoV-2 IgM and IgG enzyme-linked immunosorbent assays (ELISA).^20,21,22^ The aims of this study were to explore the performance characteristics of the Elecsys Anti-SARS-CoV-2 immunoassay and to provide detailed evidence with regard to its clinical performance, with a focus on specificity, among various prepandemic patient cohorts.

## Materials and Methods

### Study design

This retrospective, non-interventional study was conducted at five sites: one site (Innsbruck [Austria]) provided serum samples and four sites (Augsburg [Germany], Hagen [Germany], Heidelberg [Germany], and Bern [Switzerland]) provided serum and/or plasma samples and performed testing using the cobas e 801 analyser (Roche Diagnostics International Ltd, Rotkreuz, Switzerland). All samples were obtained before September 2019 and were presumed negative for SARS-CoV-2-specific antibodies. We evaluated the specificity of the Elecsys Anti-SARS-CoV-2 immunoassay in samples from blood donor screening and routine diagnostic testing, including pregnancy screening and paediatric samples. Details of the cohorts enrolled and tested across the five sites can be found in **Table 1**.

**Table 1.** Cohort details for blood donor samples and routine diagnostic specimens preceding the outbreak of SARS-CoV-2

| Group | Collection timepoint | Sample type | Site(s) | No. of samples |
| --- | --- | --- | --- | --- |
| **Group A and B** | **Pre-September 2019** | **All sample types** | **All sites** | **9575** |
| **Group A**<br>**(blood donors)** | March 2016 (flu season) | Blood donation samples (serum) | Austria (Innsbruck*) | 1048 |
|  | 2016 (no seasonal indication) | Blood donation samples (serum) | Germany (Hagen) | 2625 |
|  | Combined 2018 (flu and no flu season) | Blood donation samples (serum and plasma) | Switzerland (Bern) | 3041 |
|  | July/August 2018 (no flu season) | Blood donation samples (serum) | Switzerland (Bern) | 2003 |
|  | February/March 2018 (flu season) | Blood donation samples (plasma) | Switzerland (Bern) | 1038 |
|  | **Pre-September 2019** | **All blood donation samples** | **Austria (Innsbruck\*), Germany (Hagen) and Switzerland (Bern)** | **6714** |
| **Group B**<br>**(routine diagnostic testing)** | Pre-September 2019 | Samples from unselected routine diagnostic testing (serum) | Germany (Augsburg) | 400 |
|  | Pre-September 2019 | Samples from pregnancy screening (serum and plasma) | Germany (Augsburg) | 1498 |
|  | Pre-September 2019 | Samples from pregnancy screening (serum) | Germany (Heidelberg) | 758 |
|  | Pre-September 2019 | Samples from pregnancy screening (serum and plasma) | Germany (Augsburg and Heidelberg) | 2256 |
|  | Pre-September 2019 | Paediatric samples (serum) | Germany (Heidelberg) | 205 |
|  | **Pre-September 2019** | **All routine diagnostic testing samples (including pregnancy screening and paediatric samples)** | **Germany (Augsburg and Heidelberg)** | **2861** |

\*Samples from Innsbruck were analysed at Augsburg. SARS-CoV-2, severe acute respiratory syndrome coronavirus 2.

This study was conducted in accordance with the study protocol provided by Roche Diagnostics and in accordance with the principles of the Declaration of Helsinki. All human samples utilised were anonymised, frozen, residual samples for which no ethical approval was required. The study protocol was submitted to institutional review boards at study sites in Innsbruck (Austria) and Bern (Switzerland) prior to study initiation; ethical approval was granted for Innsbruck, and a waiver granted for Bern.

### Assay

The Elecsys Anti-SARS-CoV-2 immunoassay uses a double-antigen sandwich (DAGS) format, which requires binding of antibodies in the patient sample to two specific antigens, and as such favours the preferential detection of mature, high-affinity antibodies characteristic of the late stages of SARS-CoV-2 infection.^23,24^ The Elecsys Anti-SARS-CoV-2 immunoassay detects antibodies independent of isotype, detecting predominantly mature, high-affinity IgG but also IgA and IgM antibodies.^22^ Briefly, the immunoassay involves incubation of patient samples with biotinylated N antigen and ruthenylated N antigen, leading to the formation of DAGS complexes in samples containing the corresponding antibodies. After the addition of streptavidin-coated microparticles, solid-phase bound DAGS complexes are magnetically captured to the surface of an electrode in the detection instrument and visualised by voltage-induced chemiluminescent emission.

The test is CE mark approved and detects antibodies in serum or plasma, collected using standard sampling tubes. The duration of the analysis is approximately 18 minutes and the results are presented as a cut-off index (COI) as well as an indication as to whether a sample is ‘reactive’ or ‘non-reactive’. Results were determined automatically by the software by comparing the electrochemiluminescence signal obtained from the reaction product of the sample with the signal of the cut-off value previously obtained by calibration.

### Sample handling and data management

Anonymised (two side delinked), frozen, residual samples were thawed to room temperature and homogenised using a slow rotating system (approximately 33 rpm) or by inverting slowing five times prior to assaying to avoid the production of foam. Before testing, samples were visually inspected to check that they did not contain clots/precipitates, foam or droplets at the container walls. Assay results were obtained via instrument export files.

### Statistical analysis

An appropriate sample size was calculated using a previously published formula, to ensure α = 5% and power = 80%.^25^ Acceptance criteria required that confidence intervals (CIs) for specificity overlapped with the confidence range stated in the Elecsys Anti-SARS-CoV-2 immunoassay package insert, namely 99.65-99.91%. Point estimates and 95% CIs (two sided) for specificity were calculated based on the exact method. For the purpose of analysis, samples were grouped into those obtained from archived blood donations (Group A) and those from archived routine diagnostic specimens (Group B).

## Results

### Analysis set

From across the five sites, 9575 samples presumed negative for SARS-CoV-2 antibodies were included in the analysis; 6714 samples from archived blood donations and 2861 samples from archived routine diagnostic specimens. The diagnostic specimens included 2256 samples from pregnant women and 205 samples from paediatric patients **(Table 1)**.

### Specificity

Specificity of the Elecsys Anti-SARS-CoV-2 immunoassay for the overall sample cohort (n = 9575) and by analysis group are shown in **Table 2**. Using an assay COI of 1.0 resulted in an overall specificity of 99.85% (95% CI 99.75-99.92) in samples obtained across all five sites. Among 6714 serum and/or plasma samples from blood donors and 2861 serum and/or plasma samples from routine diagnostic samples, specificity was 99.82% (95% CI 99.69-99.91) and 99.93% (95% CI 99.75-99.99), respectively. Among 2256 samples from pregnant women, specificity was 99.91% (95% CI 99.68–99.99). Among 205 paediatric samples, specificity was 100% (95% CI 98.22-100).

**Table 2.** Summary of specificity results for the Elecsys Anti-SARS-CoV-2 immunoassay in blood donor samples and routine diagnostic specimens

| Group | Sample cohort | No. samples tested | No. samples reactive | No. samples non-reactive | Specificity, % (95% CI) |
| --- | --- | --- | --- | --- | --- |
| **Group A and B** |  | 9575 | 14 | 9561 | 99.85 (99.75–99.92) |
| **Group A (blood donors)** | Austria (Innsbruck), flu season* | 1048 | 5 | 1043 | 99.52 (98.89–99.84) |
|  | Germany (Hagen) | 2625 | 2 | 2623 | 99.92 (99.73–99.99) |
|  | Switzerland (Bern) | 3041 | 5 | 3036 | 99.84 (99.62–99.95) |
|  | Switzerland (Bern), no flu season | 2003 | 2 | 2001 | 99.90 (99.64–99.99) |
|  | Switzerland (Bern), flu season | 1038 | 3 | 1035 | 99.71 (99.16–99.94) |
|  | All | 6714 | 12 | 6702 | 99.82 (99.69–99.91) |
| **Group B (routine diagnostic testing)** | Germany (Augsburg), diagnostic routine | 400 | 0 | 400 | 100 (99.08–100) |
|  | Germany (Augsburg), pregnancy | 1498 | 2 | 1496 | 99.87 (99.52–99.98) |
|  | Germany (Heidelberg), pregnancy | 758 | 0 | 758 | 100 (99.51–100) |
|  | Germany (Augsburg and Heidelberg), pregnancy | 2256 | 2 | 2254 | 99.91 (99.68–99.99) |
|  | Germany (Heidelberg), paediatrics | 205 | 0 | 205 | 100 (98.22–100) |
|  | All | 2861 | 2 | 2859 | 99.93 (99.75–99.99) |

\*Samples from Innsbruck were analysed at Augsburg.
CI, confidence interval; SARS-CoV-2, severe acute respiratory syndrome coronavirus 2.

Across Groups A (blood donors) and B (routine diagnostic specimens), a total of 14 reactive samples were detected (Group A, n = 12; Group B, n = 2).

### COI distribution

The COI distribution across samples is shown in **Figure 1**. In total, 9561 of the presumed SARS-CoV-2 negative samples tested had a COI of <1.0; the vast majority of these had COIs <0.1 (n =9064). Only 14 samples had a COI ≥1.0 (pre-specified cut-off for reactivity).

**Figure 1.**
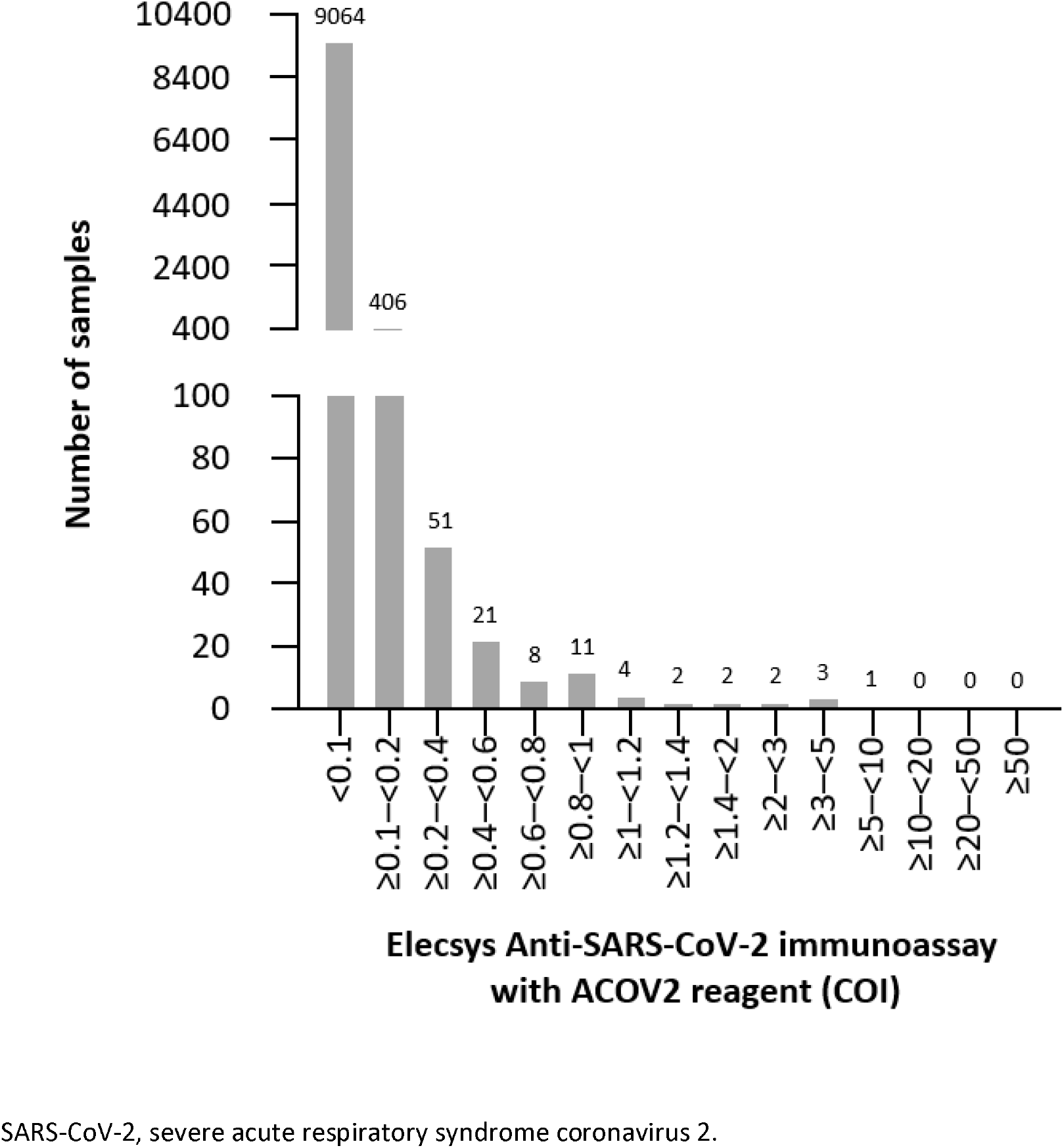
Cut-off index (COI) distribution in patient samples (n =9575)

## Discussion

Emergence of SARS-CoV-2 and the subsequent COVID-19 pandemic have resulted in an urgent, unmet need for highly specific serological tests to aid determination of past exposure to the virus, and inform seroprevalence in a given population.^26,27^ This study demonstrates that the Elecsys Anti-SARS-CoV-2 immunoassay displays very high specificity across a large cohort of blood donor and routine diagnostic samples (99.85%), supporting its use in routine testing for past exposure to SARS-CoV-2.

The very high specificity reported here for the Elecsys Anti-SARS-CoV-2 immunoassay corresponds with that reported by the manufacturer (99.78% specificity for blood donor samples; 99.81% specificity for routine diagnostic samples; 99.80% specificity overall^23^) and previous studies in samples drawn pre-COVID-19, wherein the calculated specificity for this immunoassay ranged from 98.00-99.80%.^20,21,28,29^

Importantly, the overall specificity of the Elecsys Anti-SARS-CoV-2 immunoassay determined in the present study (99.85%) is also comparable with that of other commercially available assays. These include the Wantai SARS-CoV-2 Total Antibody ELISA (100% specificity), Abbott SARS-CoV-2 IgG ELISA (99.7% specificity) and DiaSorin LIAISON® SARS-CoV-2 S1/S2 IgG ELISA (98.3% specificity).^28,30^ Specificity of the Wantai SARS-CoV-2 Total Antibody ELISA assay was evaluated using a cohort of 30 polymerase chain reaction-confirmed positive serum samples and 82 control serum samples, whereas specificities of the Abbott SARS-CoV-2 IgG ELISA and DiaSorin LIAISON® SARS-CoV-2 S1/S2 IgG ELISA were determined using 1554 serum samples collected pre-COVID-19, and therefore presumed negative for SARS-CoV-2.^28,30^ Higher specificity was observed for the Elecsys Anti-SARS-CoV-2 immunoassay compared with the EuroImmun Anti-SARS-CoV-2 IgA and IgG ELISAs, for which 93% and 96% specificity, respectively, were previously observed.^30^ The relatively lower specificities measured for these two assays could be attributed to cross-reactivity with antibodies to adenoviruses and other human coronaviruses,^30^ which may result in false positive/negative results. Encouragingly, Muench et al. recently demonstrated that among 80 samples tested for crossreactivity with common cold or endemic coronavirus panels using the Elecsys Anti-SARS-CoV-2 immunoassay, none were reactive.^21^ The present analyses demonstrate a marginal decrease in specificity for the Elecsys Anti-SARS-CoV-2 immunoassay in blood donor samples taken during flu season (Bern, 99.71% [95% CI 99.16-99.94]; Innsbruck, 99.52% [95% CI 98.89-99.84]) compared with samples drawn outside of flu season (Bern, 99.90% [95% CI 99.64-99.99]), which may be a result of exposure to other immune stimuli in the form of seasonal flu. This difference was not statistically significant, however, and the Elecsys Anti-SARS-CoV-2 immunoassay displayed very high specificity in both groups.

The Elecsys Anti-SARS-CoV-2 immunoassay also demonstrated very high specificity in samples from pregnant women (99.91%). Accurate serological assays for SARS-CoV-2 are especially important for use in pregnant women, for whom altered immune status may increase susceptibility to respiratory virus infections, and may therefore have implications for clinical management in this setting.^31^ At present, there are limited data available on the performance of other commercially available serological assays for SARS-CoV-2 in pregnant women. Rushworth et al. conducted a performance evaluation of the Mount Sinai SARS-CoV-2 IgG antibody ELISA, and observed no false positive results in a cohort of negative control samples (n = 50) drawn from presumed healthy pregnant women pre-COVID-19.^32^ In the present study, two reactive results were observed in prepandemic samples taken from pregnant women using the Elecsys Anti-SARS-CoV-2 immunoassay, which could be the result of an unknown cross-reactant.

Excellent specificity was observed for this immunoassay in a paediatric cohort (100% specificity), indicating strong performance in this setting; however, it should be noted that the statistical significance of these results is not very powerful due to the relatively small cohort of samples used (n = 205). Further investigation into and validation of other commercially available assays within this setting is warranted.

A major strength of this study is the large cohort of presumed SARS-CoV-2-negative serum and plasma samples (n = 9575) used to evaluate specificity of the Elecsys Anti-SARS-CoV-2 immunoassay, across multiple sites in different countries, ensuring that our data are reliable and can be used to form robust comparisons with immunoassay performance data from other studies. To our knowledge, the present study includes the largest cohort of samples from pregnant women used to date to evaluate the performance of a serological assay for SARS-CoV-2 with regard to specificity. Existing literature on the performance of commercially available assays for detection of antibodies to SARS-CoV-2 is limited in pregnant women and paediatric populations; thus, additional research in these groups is warranted to further inform clinical decision-making.

## Conclusion

The performance of SARS-CoV-2 antibody assays in general is of high importance for public health and may affect political decision-making in pandemic management. This study generated additional data on the performance of the Elecsys Anti-SARS-CoV-2 immunoassay and provided broader evidence on the very high specificity of the assay, including in cohorts of pregnant women and paediatric populations.

## Data Availability

Qualified researchers may request access to individual patient level data through the clinical study data request platform (https://vivli.org/). Further details on Roche's criteria for eligible studies are available here: https://vivli.org/members/ourmembers/. For further details on Roche's Global Policy on the Sharing of Clinical Information and how to request access to related clinical study documents, see here: https://www.roche.com/research_and_development/who_we_are_how_we_work/clinical_trials/our_commitment_to_data_sharing.htm

## Acknowledgements

The authors thank: Annelies Mühlbacher (University Hospital LKH) for provision of pre-COVID-19 pandemic blood donation samples, which were tested in Augsburg; Peter Gowland and Caroline Tinguley (Interregionale Blood Transfusion Swiss Red Cross) for involvement in preparation of prepandemic samples and laboratory work; Florina Langen (Roche Diagnostics) for study conceptualisation and drafting of the protocol, study management, interpretation of analysis and further critical input; Christopher Rank (Roche Diagnostics) for database generation and data validation, authorship of the statistical analysis plan and involvement in the formal analysis; Sigrid Reichhuber, Janina Edion and Yvonne Knack (Roche Diagnostics) for management of investigation sites, data acquisition and study monitoring.

Third-party medical writing support, under the direction of the authors, was provided by Chloe Fletcher (Gardiner-Caldwell Communications, Macclesfield, UK) and was funded by Roche Diagnostics International Ltd, Rotkreuz, Switzerland.

COBAS, COBAS E and ELECSYS are trademarks of Roche.

## Conflicts of interest

- **Elena Riester:** received speaker’s honorarium from Roche.
- **Beda Krieter:** no conflicts of interest to disclose.
- **Peter Findeisen:** no conflicts of interest to disclose.
- **Michael Laimighofer:** employee of Roche Diagnostics GmbH.
- **Kathrin Schoenfeld:** employee of Roche Diagnostics GmbH; owner of shares in Roche.
- **Tina Laengin:** employee of Roche Diagnostics GmbH.
- **Christoph Niederhauser:** no conflicts of interest to disclose.

## Funding

This study was funded by Roche Diagnostics GmbH (Mannheim, Germany).

## Required information for submission system

*Ethical guidelines*

*Research reporting guidelines*

Please see separate STARD checklist.

*Data availability statement*

